# Interrogating the complex relationship between Alzheimer’s disease and blood pressure: two instrumental variable approaches to focus on pre-clinical stages of Alzheimer’s disease in UK Biobank

**DOI:** 10.1101/2024.10.17.24315654

**Authors:** Jennifer C Palmer, Emma Hart, Emma Anderson, Seth Love, Deborah A Lawlor

## Abstract

**Background and Aims:** Evidence suggests there may be a bidirectional relationship between high blood pressure (BP) and Alzheimer’s disease (AD). It is hypothesised that this is due to cerebral changes during pre-clinical AD that cause elevation of systemic BP. We aimed to test this by exploring the effect of risk of pre-clinical AD on blood pressure.

**Methods:** We used data from the UK Biobank, including adults without prevalent or incident (within first 5-years of follow-up) clinical AD (N = 501,420, mean age 56.6, SD 8 years). We used two instrumental variables, an age-weighted parental dementia instrument score and a participant genetic instrument score, that are vulnerable to differing biases, to instrument risk of pre-clinical AD (the exposure). We tested the association of both instrument scores with systolic BP (SBP), diastolic BP (DBP) and hypertension. Sensitivity analyses were undertaken to explore different biases.

**Results:** Both the higher parental dementia instrument and participant genetic instrument score were associated with higher mean SBP (difference in mean SBP mmHg per 1SD higher score: 0.12, 95% CI: 0.06 to 0.17, and 0.07, 95% CI: 0.00 to 0.13, respectively) but not DBP. Sensitivity analyses were largely consistent with these findings.

**Conclusions:** Our findings provide preliminary evidence that pre-clinical AD increases SBP. Further research is required to determine whether this increase in SBP is due to increased cerebrovascular resistance in pre-clinical AD. Obtaining a better understanding of the changing relationship with BP at different stages of AD may enable effective optimisation and targeting of therapies.

## Introduction

Alzheimer’s disease is often divided into three stages based on the clinical presentation: the pre-clinical stage, characterised by normal cognition; the prodromal stage, characterised by mild cognitive impairment; and the dementia stage, where there is functional cognitive impairment.^1^ Although Alzheimer’s disease dementia usually occurs in later life, there are early disease processes in the brain, such as a decline in cerebral blood flow, that precede clinical presentation by up to 20 years before diagnosis.^2–5^

Hypertension is the leading preventable risk factor for cardiovascular disease and mortality worldwide. Hypertension affects 1 in 3 adults worldwide, and globally nearly half of people with hypertension are unaware of their condition.^6^ Often, the cause of elevated blood pressure (BP) is not identified, but it may stem from reduced perfusion to any of several major organs, namely kidney, lungs and, importantly, the brain.^7^

Observational evidence suggests that hypertension which develops during mid-life (45-68 years) is associated with increased risk of Alzheimer’s disease later in life.^8–10^ Some studies have found that having high BP later in life (>65 years) is associated with lower risk of Alzheimer’s disease.^11,12^ The results of observational studies may be exaggerated by residual confounding. They may also be underestimated by survivor bias, given that those with higher BP from earlier in life are more likely to die in a given time period compared to those with lower BP, or those whose BP rises only in later life. Randomized controlled trials of anti-hypertensives for the treatment of Alzheimer’s disease have been inconsistent, often showing no significant risk reduction.^13,14^

An instrumental variable is a variable that is associated with the exposure of interest and is only associated with the outcome of interest through its association with the exposure. Instrumental variable analyses can be used to estimate a causal effect between an exposure and outcome, and may be less prone to confounding than using the exposure directly, if three strong assumptions outlined in Table 1 and Figure 1 are met.^15^ Mendelian randomization is a method that commonly uses genetic variants as instrumental variables to determine the unconfounded effect of potential risk factors on outcomes.^16^ Mendelian randomization studies to date have explored the potential effect of higher BP on Alzheimer’s disease and have reported differing results, including evidence for higher BP resulting in lower risk of Alzheimer’s disease, higher risk and no evidence of effect.^17–25^ These differences may be explained by between-study differences in instrument strength, and in the extent to which they explored survival bias and bias due to horizontal pleiotropy. We did not identify any specific Mendelian Randomization study of the effect of Alzheimer’s disease on blood pressure. However, a Mendelian randomization phenome-wide association study exploring the potential effect of Alzheimer’s disease on various health outcomes suggested that genetic liability to Alzheimer’s disease had an age-related effect on SBP, potentially resulting in higher SBP in younger participants (aged 39-53) but not older participants (53-72 years).^26^ No sensitivity analyses were undertaken for this potential effect.

**Table 1.** The assumptions underpinning instrumental variable analyses, and how they have been met or violated in this study.

| *Instrumental variable (IV)*<br>*assumption* | *How assumption is met or violated* |  |
| --- | --- | --- |
|  | **with PRS as instrument** | **with GRS as instrument** |
| **IV1. IV is statistically strongly associated with the exposure of interest in the relevant population** | Having one or both parent(s) with clinical dementia is associated with increased risk of developing Alzheimer's disease themselves. | Genetic variants associated at genome-wide significance ($p < 0.05 \times 10^{-8}$) to AD in a chosen pre-existing AD GWAS<sup>37</sup> were used. |
| **IV2. There is no confounding between the IV and outcome** | This is likely to be violated by a confounding path between parental and participant education, ethnicity, smoking, physical activity, BMI and alcohol. We adjusted for parental measures of these. Residual confounding due to lack of parental measures and potential imprecise measures e.g. of self-report smoking, alcohol add physical activity could bias results away from the null. | As genetic variation is fixed at conception, common confounders, such as education, ethnicity, BMI etc. cannot confound the participant genetic instrument score. Confounding can occur where there are subgroups of different populations (e.g. groups from different ancestral backgrounds) who have different allele frequencies of the AD related genetic variants, and independently of that have different BP distributions. We limited our analyses to self-declared white European ethnicity to mitigate against that and adjusted genetic analyses for ancestral principal components. |
| **IV3. The IV does not influence the outcome by any other path than via the exposure of interest** | If parental dementia affected offspring adult blood pressure via a direct path, in addition to a possible effect mediated by offspring pre-clinical dementia this assumption would be violated. We cannot think of such a path and so assume this is not violated. | A pathway which can lead to an association between the genetic variants and blood pressure is through horizontal pleiotropy ($C_2$ in Figure 1B). This will be explored with sensitivity analysis, and steps will be taken to minimise. |
| *Additional two-sample MR assumptions* |  |  |
| **MR1. The genetic IV-exposure association study sample and the genetic-outcome association sample are from the same underlying population and have little or no overlap** |  | Both samples were of White European ancestry only and we selected the largest and most appropriate AD GWAS that did not include UK biobank to ensure the two samples were largely independent (non-overlapping). |
| **MR2. The genetic variants are harmonised (same reference allele) across the two samples** |  | Variants will be harmonised, and harmonisation checked by comparing allele frequencies of the reference and effect alleles. |

**Figure 1:**
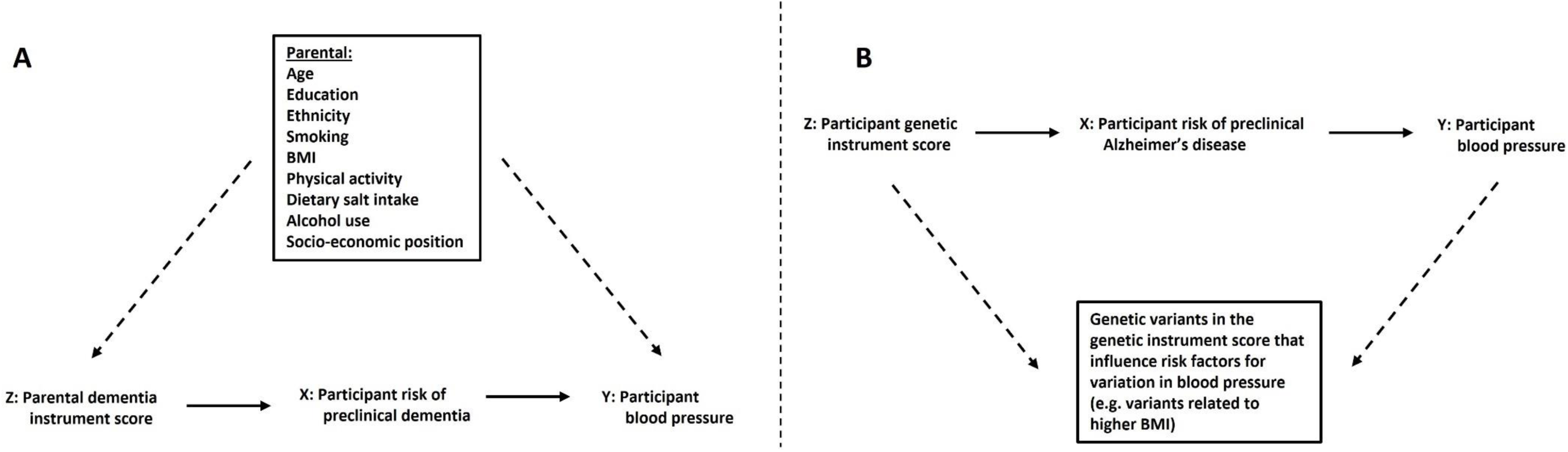
Directed Acyclic graphs depicting the hypothesised causal pathways using (A) parental dementia instrument score as an instrumental variable and (B) participant genetic instrument score as an instrumental variable for risk of pre-clinical Alzheimer’s disease. Directed Acyclic graphs (DAGs) place arrows between a variable at the base of the arrow and one at the head (point of the arrow) when there is a known or plausible causal effect of the variable at the base on the variable at the top. DAGs are useful for illustrating analysis assumptions and here we use them to show that our two instrumental variables have different key sources of bias). Instrumental variable assumptions are described in **Table 1** and summarised in relation to this figure here: 1. Relevance assumption – that there is a statistically robust association between the instrument and exposure in the relevant population; shown here by the solid arrow from the parental dementia instrument score (in A) and from the participant genetic instrument score (in B) to risk of pre-clinical Alzheimer’s disease. 2. Independence assumption – that there is no confounding between the instrumental variable and the outcome; because we believe this is plausibly violated for the parental dementia instrument score, we have used dashed arrows to indicate which factors plausibly violate this assumption. For the participant genetic instrument score (in B) there are no arrows suggesting confounding between the participant genetic instrument score and variation in blood pressure, since it is not plausible for factors such as age, sex, BMI, etc. to influence genetic variation. 3. Exclusion restriction criteria – that the association of the instrument with the outcome is only due to the relation of the instrument with the exposure (i.e. there are no direct paths from the instrument to the outcome that are independent of the exposure. Because we believe it is plausible that genetic variants related to Alzheimer’s disease could relate to other risk factors for variation in blood pressure, we have dashed arrows to show this (in B), which are not present in the parental dementia instrument score graph (A). Conventional DAG notation is used: Z: instrumental variable; X: exposure being instrumented; Y Outcome

One hypothesis that could explain some of these inconsistencies is that during the pre-clinical period of Alzheimer’s disease, reduced cerebral blood flow can cause an elevation in peripheral BP in order to protect healthy cerebral perfusion (analogous to the development of renovascular hypertension or pulmonary hypertension in people with increased renal or pulmonary vascular resistance) which benefits cognitive function.^27^ However, prolonged elevation of BP over many years leads to structural vascular changes including arteriosclerosis, causing secondary hypoperfusion and blood-brain barrier breakdown, which accelerate the progression of cognitive decline.^12^ Thus, it has been hypothesized that there might be a bi-directional relationship between high BP and Alzheimer’s disease.^27^

Evidence for the bi-directional Alzheimer’s disease-BP hypothesis has mainly come from biochemical measurements in post-mortem brain tissue,^28^ as summarized by Love and Miners.^27^ Causal evidence for this hypothesis in human populations is limited. Observational studies to date are mostly designed to try to capture the risk of prolonged elevation of BP on Alzheimer’s disease. Interpretation of results from traditional observational approaches is challenging as most in fact are capturing the risk associations in both directions (e.g. particularly when cohorts span a wide age range and follow-up period).

The aim of this study was to test the bi-directional Alzheimer’s disease-BP hypothesis in the absence of good measures of cerebral blood flow in large human epidemiological studies. We address this aim by determining the association of two different instrumental variables for pre-clinical Alzheimer’s disease with subsequent BP, in humans not, or not yet, experiencing prodromal or clinical Alzheimer’s disease. The first instrument was a ‘parental dementia instrument score’, with participants given a score based on their self-report of whether neither, one or two of their parents had been diagnosed with all-cause dementia, adjusted for parental age to reflect our confidence in likelihood of dementia diagnosis (ie younger parents may have gone on to develop dementia had data been collected later or had they lived longer). The second instrument was a ‘participant genetic instrument score’, which was constructed by generating a weighted allele score of alleles in Single Nuclear Polymorphism (SNPs) that in a large genome-wide association study (GWAS) are significantly associated with odds of Alzheimer’s disease.

The value of using these two different instrumental variables is that they have different and unrelated key sources of bias, so if both are associated with higher BP that would provide stronger evidence for the hypothesis than if we had results from just one of them.^29^ The key potential source of bias in the parental dementia instrument score is confounding between that instrument and BP, whereas the key potential bias in the participant genetic instrument score is unbalanced horizontal pleiotropy resulting in a direct path from the instrument to the outcome, independent of Alzheimer’s disease (Table 1 and Figure 1).

## Methods

### Study population and analyses samples

We used data from the UK Biobank, a large prospective cohort study that recruited 503,325 adults (5.5% of those invited) aged between 40 and 69 years and recruited from across the UK between 2006 and 2010.^30^ Of the 503,325, 912 were excluded because they had withdrawn their data from the study. All participants had provided written informed consent, approved by the Northwest Multicentre Research Ethics Committee. As data were deidentified, this study did not require additional ethics committee approval. We published all methods of data preparation, validation and analysis online *a priori* prior to running analyses^31^ and this study followed the Strengthening the Reporting of Observational Studies in Epidemiology (STROBE) reporting guidelines.^32^

We excluded 993 participants who had been diagnosed with prevalent dementia, or incident dementia within the first five years of recruitment, to avoid reverse causality. Prevalent dementia (n=89) was obtained from the base-line self-completed diseases questionnaire. Information on incident cases (n=904) was obtained from available linked primary care records and hospital episode statistics (see Supplementary Table 1 for ICD-9 and ICD-10 codes used for exclusion). Of the remaining 501,420 participants, 445,911 had a valid parental dementia instrument score, and 336,946 had a valid participant genetic instrument score (excluding those that failed quality control analyses run using a previously developed QC pipeline,^33^ as well as non-European participants to avoid potential confounding by population stratification). Missing outcome and confounder data were less than 0.5%, other than education which was missing for 1.5% participants (Table 2 and Supplementary Table 2).

**Table 2.** Description of participants included in each analysis.

| Observed variable | All eligible participants<br>(PDIS cohort) |  | Complete data, PDIS<br>analysis cohort** |  | Complete data, PGIS<br>analysis cohort*** |  |
| --- | --- | --- | --- | --- | --- | --- |
|  | n (%) | N* | n (%) | N | n (%) | n |
| **Categorical** |  |  |  |  |  |  |
| Women | 272,930 (54) | 501,420 | 239,901 (55) | 433,764 | 180,776 (54) | 336,295 |
| Education, highest qualification CSE/O-level/GCSE or equiv. | 217,047 (43) | 492,187 | 185,224 (43) | 433,764 | 148,852 (44) | 333,212 |
| Ethnic minority groups | 26,968 (5) | 498,659 | 20,254 (5) | 433,764 | 0 (0) | NA |
| Current smoker | 52,845 (10) | 498,500 | 43,238 (10) | 433,764 | 33,276 (10) | 334,827 |
| High salt diet (always add salt to food) | 24,363 (5) | 500,317 | 19,931 (5) | 433,764 | 14,420 (4.3) | 335,969 |
| Use alcohol daily/almost daily | 101,567 (20) | 499,944 | 89,742 (21) | 433,764 | 72,029 (21) | 335,772 |
| Do 10 mins or more of vigorous physical activity 3 or more times per week | 181,982 (36) | 501,420 | 153,852 (36) | 433,764 | 118,739 (35) | 336,005 |
| **Continuous** | **mean (sd)** | **N** | **mean (sd)** | **N** | **mean (sd)** | **N** |
| Age in years | 56.5 (8.1) | 501,420 | 56.4 (8.0) | 433,764 | 56.9 (8.0) | 336,295 |
| SES by TDI | -1.3 (3.1) | 500,798 | -1.4 (3.0) | 433,764 | -1.6 (2.9) | 336,005 |
| BMI | 27.4 (4.8) | 498,673 | 27.3 (4.8) | 433,764 | 27.4 (4.8) | 335,339 |

PDIS = parental dementia instrument score; PGIS = participant genetic instrument score; SES = socioeconomic status; TDI = Townsend deprivation index; SBP = systolic blood pressure
\* with available data for that variable
\*\* complete data for PDIS-SBP associations and fully adjusted model
\*\*\* complete data for PGIS-SBP associations and additional exclusions

### Exposure instrumental variables

We used parental diagnosis of all-cause dementia as one of our instrumental variables for risk of Alzheimer’s disease in offspring. Information on parental all-cause dementia was obtained from the baseline self-completed electronic questionnaire. Participants were asked to report whether their mother or father had ever suffered from Alzheimer’s disease or dementia, and to report each parent’s current age if still alive, or age at death if not. From this information we created a weighted instrumental variable for risk of pre-clinical dementia with an approach similar to that used in a previous study.^34^ This weighting means that each parent diagnosed with dementia contributed one full unit to the participant’s instrument score, while each parent without dementia contributed less than one unit, inversely proportional to their age, reflecting our confidence in their not having dementia, such that weight=(100-age)/100 (age 100 is approximately the 95^th^ percentile for life expectancy in developed countries). We used a cap at age 68 so that any parent aged 68 years or younger at the point of information collection, without diagnosed dementia, contributed the same level of (low) weight (because dementia diagnosis rises steeply with increasing age above age 68, but remains very low under age 68).^35,36^ For example, if a parent reached 100 without a dementia diagnosis, we are confident in giving 0 weight to the instrumental variable score for pre-clinical dementia. However, if a parent was 68 or younger without a dementia diagnosis, we could not be confident in giving 0 weight to the instrumental variable score for pre-clinical dementia, so gave a higher contribution of 0.32.

We also used genetic liability to Alzheimer’s disease (SNPs) as an instrumental variable for risk of pre-clinical Alzheimer’s disease. This instrumental variable was generated by creating a weighted participant genetic instrument score of 32 genome-wide significant (p <= 0.05%10^−8^) and independent SNPs selected from a previously published GWAS^37^ and weighted by the magnitude of the association with Alzheimer’s disease from that GWAS. We chose this Alzheimer’s disease GWAS for the following reasons: 1) It has close case-control matching by mean age of assessment (72.9 years for cases and 72.4 years for controls); 2) 100% of cases and 86.2% of controls had undergone clinical or pathological assessment; and 3) it did not contain data from UK Biobank, which we use for our main analyses. Within groups of correlated SNPs we selected the SNP with the strongest Alzheimer’s disease association to remain in the score. We calculated a participant genetic instrument score for pre-clinical Alzheimer’s disease by multiplying the number of effect alleles for each participant in UK Biobank by the weight, then summing across all 32 SNPs.

### Outcomes

Two BP measurements were taken by trained staff on each participant, usually using an automated device (Omron Digital BP monitor), as part of the in-person assessment. We used the mean of both automated measurements (or manual readings if automated readings were missing) of systolic BP (SBP) from the baseline assessment to generate a continuous SBP variable. If only one reading was recorded at baseline assessment, we took this as the mean SBP. If both readings were missing at baseline assessment, we used the mean of both readings at a subsequent visit instead. We used the same procedure to generate the continuous diastolic BP (DBP) variable.

We generated a binary outcome variable of hypertension, by combining those who self-reported essential hypertension, with those taking an anti-hypertensive medication (detailed below), as well as those with an SBP measurement above 140 mmHg or those with a DBP measurement above 90 mmHg. This identified those with unknown, undiagnosed hypertension, as well as those with known hypertension. A Venn diagram showing the overlap between self-report hypertension, taking anti-hypertensive medications, high SBP and high DBP is provided in Supplementary Figure 1.

For any participants who were on antihypertensive medication, or on medications that were prescribed for other conditions, such as heart failure, but that also reduce BP, we applied +15mmHg to SBP and +10mmHg to DBP, which are conservative estimates of average BP lowering by antihypertensive medications.^38^ Information on use of medications was obtained at the baseline assessment. Participants were asked to bring along all packets of prescribed medication they usually took (not purchased over-the-counter and not prescribed as a one-off) and these were recorded by nurses.

### Confounders and covariables

The parental dementia instrument score could violate the assumption of no confounding between the instrumental variable and outcome, whereas confounding is not plausible for the participant genetic instrument score (Figure 1, Table 1). We considered the following to be confounders for the parental dementia instrument score analyses based on their known or plausible influence on Alzheimer’s disease and BP: age; sex; education; ethnicity; smoking; BMI; physical activity; dietary salt intake; alcohol use and socio-economic position. Ideally, we would have had data on parental and participant measures of these. We only had measures on the participants and demonstrate in Figure 1 why we consider controlling for participant confounders should block the confounding path. Details of how these variables were assessed are reported in the Supplementary Methods.

### Statistical analyses

We summarised baseline characteristics for all eligible participants and the data for both analysis cohorts and presented as percentages for categorical variables and means with SDs for normally distributed continuous variables.

We used multivariable linear regression to determine the association of each instrument score with SBP and DBP and logistic regression for their association with hypertension. With parental dementia instrument score, we added each confounder one-by-one and assessed the standard error for evidence of multicollinearity or missing data effects on standard error. We then corrected BP for anti-hypertensive medication use. Our *a priori* assumption was that for parental dementia instrument score, our final model would be the full confounder-adjusted model with BP correction. With participant genetic instrument score as instrument, our *a priori* assumption was that genetic variants would not be associated with potentially confounding factors (but we checked this using linear or logistic regression). However, as the GWAS estimates used in our participant genetic instrument score had been adjusted for age and sex, for consistency our main model was adjusted for genetic principal components, age and sex, with BP correction for anti-hypertensive medication use.

For the participant genetic instrument score, we explored between-SNP heterogeneity, which if present could indicate violation of the assumption that there are no indirect paths from the participant genetic instrument score to BP independent of pre-clinical Alzheimer’s disease. We also performed leave-one-out analyses of SNPs in the participant genetic instrument score to assess any influential outliers.

## Results

### Descriptive analyses

The distributions of all potential confounding factors were very similar in the analysis cohort and in the complete data for both the parental dementia instrument score and participant genetic instrument score cohort (Table 2). Participants contributing to parental dementia instrument score analyses had mean age 56.4, SD 8.0 years and to participant genetic instrument score analyses mean age 56.9, SD 8.0.

### Instrumental variable associations with blood pressure

In the analysis samples for each instrument, and focusing primarily on the fully adjusted modes, we found evidence to suggest that risk of pre-clinical Alzheimer’s disease, as instrumented both by higher levels of the parental dementia instrument score and participant genetic instrument score, carried a liability to higher mean SBP (Figure 2), but no strong evidence with either instrument associating risk of pre-clinical Alzheimer’s disease to DBP (Figure 3). Risk of pre-clinical Alzheimer’s disease as instrumented by higher levels of the parental dementia instrument score was also associated with hypertension, with a weaker association when instrumented by the participant genetic instrument score (Figure 4). With the parental dementia instrument score there was a change in direction for the association with SBP following adjustment for age, from a reduction in SBP (difference in mean −0.22 mmHg) per standard deviation higher in parental dementia instrument score for the unadjusted and uncorrected analysis (Figure 1A, Model 2) to an increase in SBP (mean +0.18 mmHg) per higher standard deviation in parental dementia instrument score after adjustment for participant age at BP measurement (with a similar change in direction for hypertension). With the participant genetic instrument score, point estimates for all three of SBP, DBP and hypertension did not change with correction for use of antihypertensives, adjustment of age and sex or adjustment for principal components (Figures 2-4). In the leave-one-out sensitivity analyses for the participant genetic instrument score, removal of each SNP one at a time produced results that were consistent with the result that included all SNPs suggesting that there were no individual SNPs in the participant genetic instrument score that substantially influenced results (Supplementary Figures 2-4).

**Figure 2.**
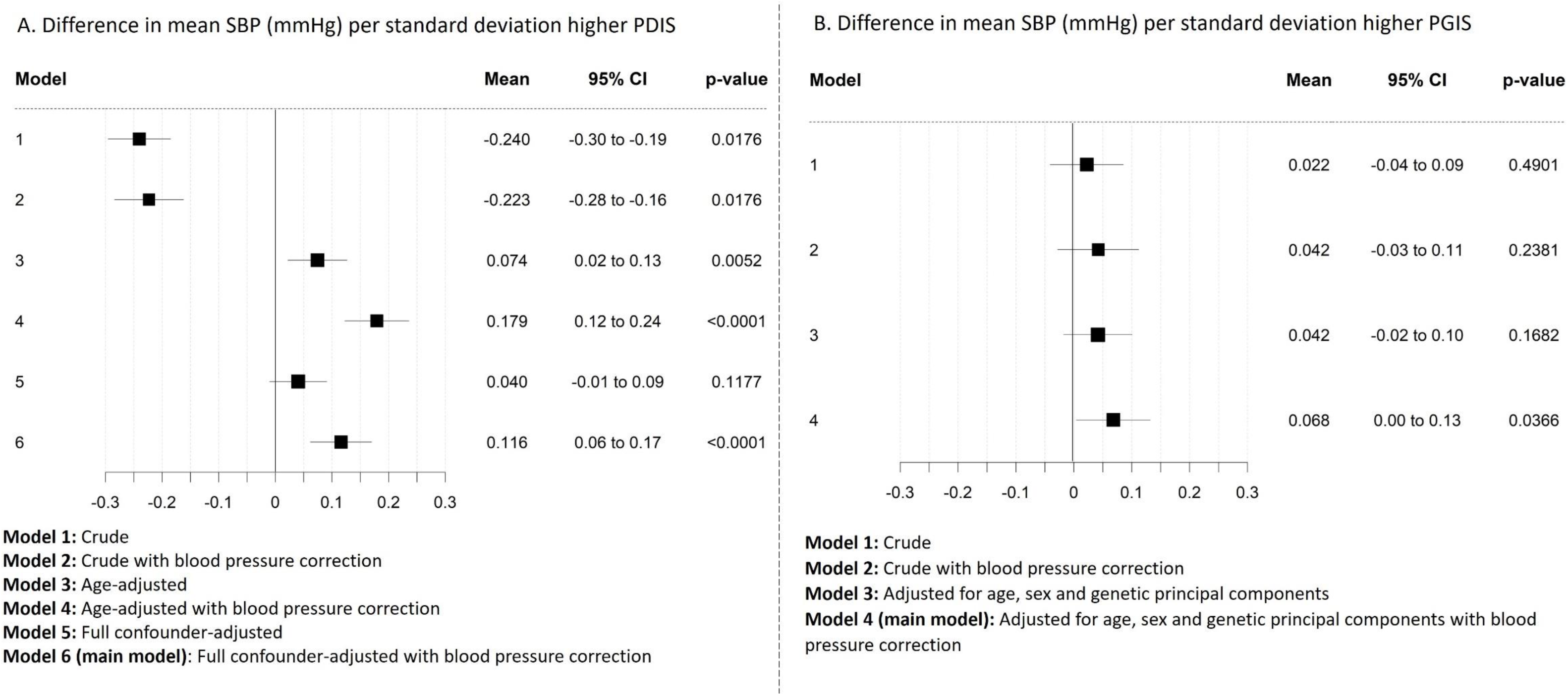
Difference in mean systolic blood pressure (SBP, mmHg) (black squares), and 95% confidence intervals (horizontal lines) per standard deviation higher parental dementia instrument score (PDIS, A) as instrumental variable for risk of pre-clinical Alzheimer’s disease and participant genetic instrument score (PGIS, B) as instrumental variable for risk of pre-clinical Alzheimer’s disease. Each model (from crude to fully adjusted) is shown to indicate the differing effects of confounding with each instrument (due to each being vulnerable to differing biases (see main text for details).

**Figure 3.**
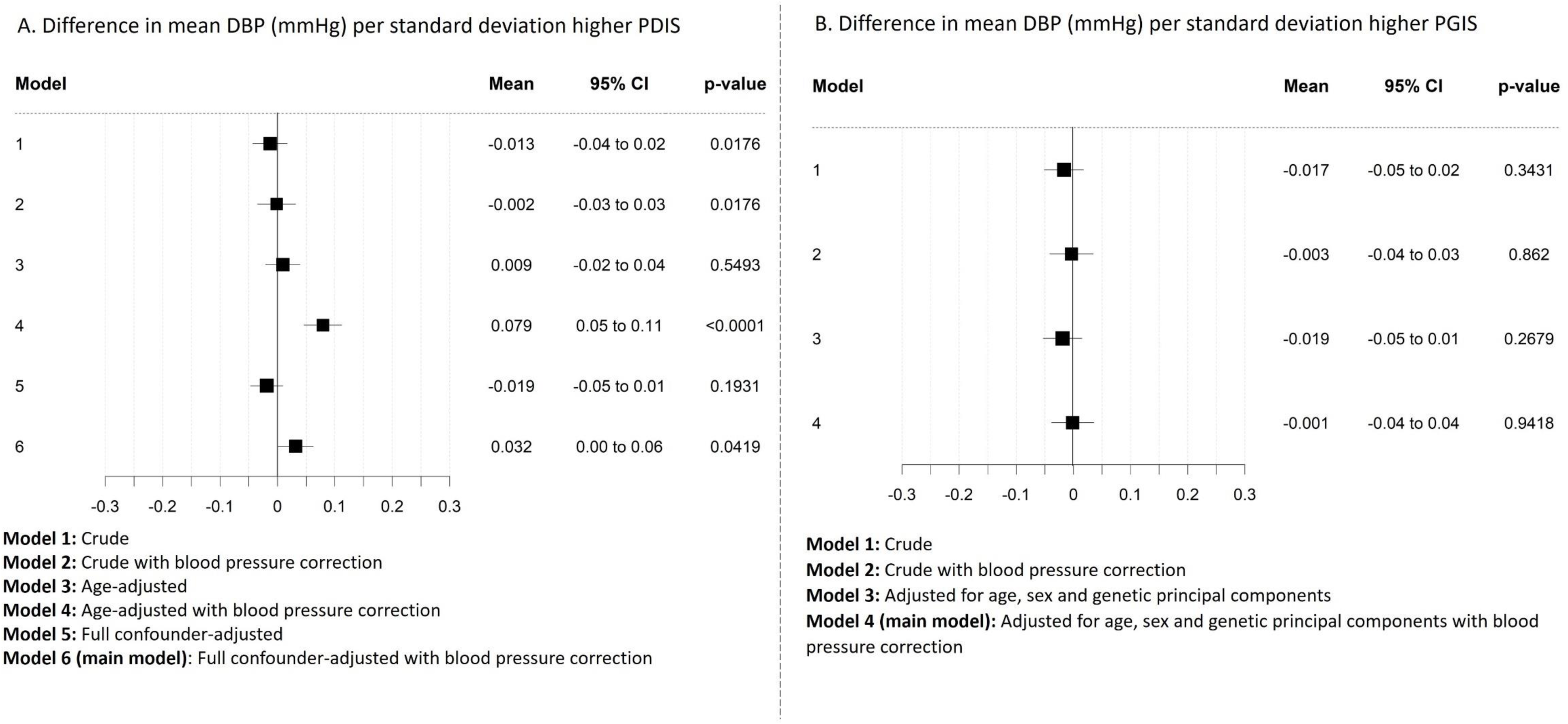
Difference in mean diastolic blood pressure (DBP, mmHg) (black squares), and 95% confidence intervals (horizontal lines) per standard deviation higher parental dementia instrument score (PDIS, A) as instrumental variable for risk of pre-clinical Alzheimer’s disease and participant genetic instrument score (PGIS, B) as instrumental variable for risk of pre-clinical Alzheimer’s disease. Each model (from crude to fully adjusted) is shown to indicate the differing effects of confounding with each instrument (due to each being vulnerable to differing biases (see main text for details).

**Figure 4.**
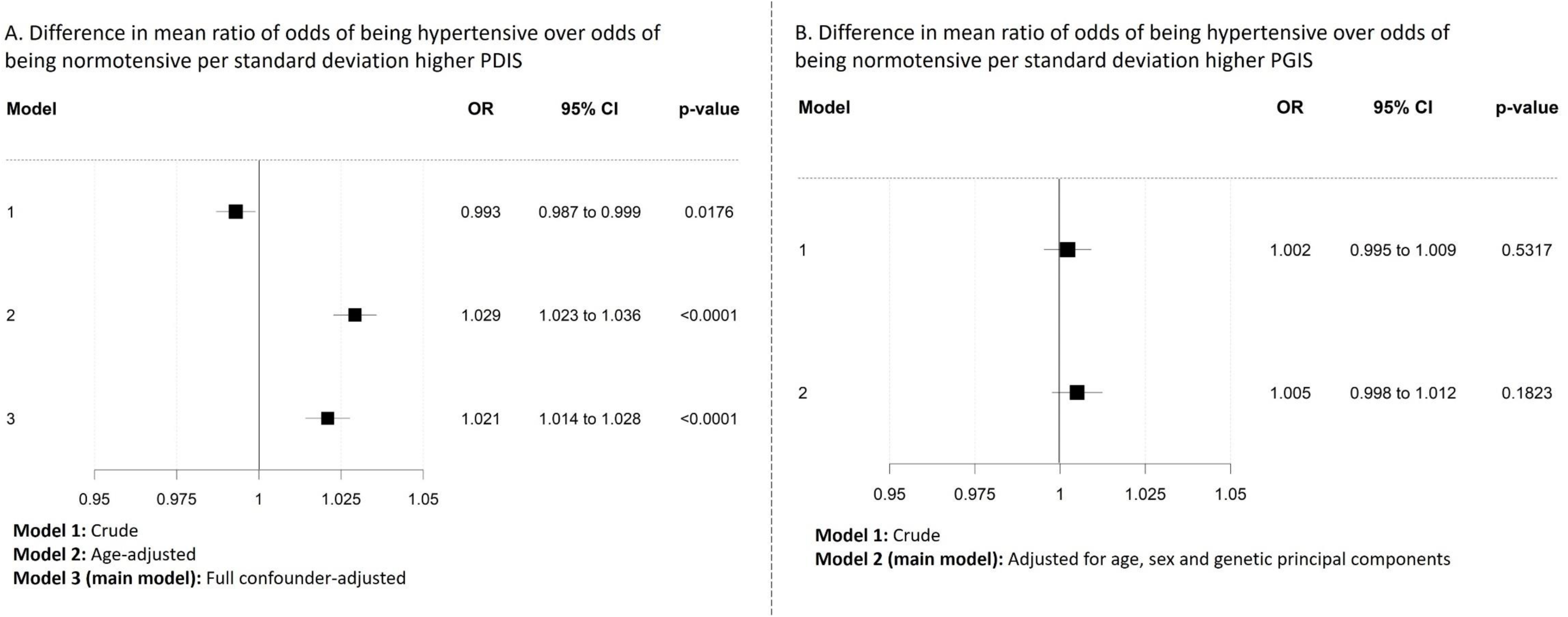
Difference in mean ratio of odds of being hypertensive over odds of being normotensive (odds ratio, black squares), and 95% confidence intervals (horizontal lines) per standard deviation higher parental dementia instrument score (PDIS, A) as instrumental variable for risk of pre-clinical Alzheimer’s disease and participant genetic instrument score (PGIS, B) as instrumental variable for risk of pre-clinical Alzheimer’s disease. Each model (from crude to fully adjusted) is shown to indicate the differing effects of confounding with each instrument (due to each being vulnerable to differing biases (see main text for details).

## Discussion

In this large human study, we have found, preliminary evidence that SBP (but not DBP) is elevated in people at risk of pre-clinical Alzheimer’s disease but without any clinical evidence of cognitive impairment or dementia. To the best of our knowledge this is the first study to explore the hypothesis that risk of pre-clinical Alzheimer’s disease results in higher blood pressure.

We used two complementary instrumental variables of a risk of pre-clinical Alzheimer’s disease. These complement each other by having different key sources of bias. The key source of bias for the parental dementia instrument score is confounding due to age, smoking, body mass index and other factors that would cause parental dementia and variation in offspring BP. By contrast the genetic instrument score is less likely to be influenced by such confounders as genetic variation is fixed at conception and cannot be changed by someone’s age, smoking, body mass or similar factors. The change in direction of associations for the parental dementia instrumental score with adjustment for age, with no change in the genetic instrument score with age, sex or principal component adjustment, supports the importance of confounding with the parental dementia instrument score, particularly by age. By taking parental age into account in our parental instrument score, it is less prone to survival bias than a simple ‘count’ score of whether a parent had been diagnosed with dementia. If using a simple count, the accuracy of the score is associated with parental age (the older the parents the more accurate the score), as for a dementia diagnosis they will have had to reach that age. As BP is also heritable, and increases mortality risk, we would expect an inverse association between BP and a simple parental count score, largely driven by survival bias due to differential misclassification of non-dementia (that would have been scored as parental dementia had parents lived longer or participants been assessed later). Our parental instrument score accounts for age, to represent our certainty of correct classification of non-dementia, and thus is less prone, although still not possible to be free from, survivor bias. The key source of bias for the participant genetic instrument score is horizontal pleiotropy, which we cannot rule out here, though our leave-one-out analyses suggest no single SNP has a major effect on the main result, and the consistency across results after removing each one individually provides some support for horizontal pleiotropy not having a major effect. However, the consistency of direction of association across the two instruments, with higher mean SBP and higher odds of hypertension demonstrated in both, despite their differences in biases, supports the hypothesis that a risk of pre-clinical Alzheimer’s disease causes a rise in SBP.

Our results have relevance to previous studies suggesting possible complex relations between BP and Alzheimer’s disease, and address many of the limitations of those previous studies. The large sample size enabled us to exclude participants with prevalent and incident dementia in the first five-years of follow-up, so that associations reflect pre-clinical disease and importantly are unlikely to be driven by established dementia or cognitive impairment as a consequence of prolonged elevation of BP.

### Mechanisms: why is SBP elevated in people with pre-clinical Alzheimer’s disease?

We found that risk of pre-clinical Alzheimer’s disease, as measured using both a parental dementia instrument score and a participant genetic instrument score, associates with higher peripheral SBP (but not DBP). We hypothesis that this is due to very early reductions in cerebral perfusion. This is consistent with the physiological and laboratory studies informing each part of the full hypothesis: cerebral blood flow is reduced early in Alzheimer’s disease,^2,3^ for example, decreases in grey-matter CBF were documented in young individuals (aged 19-32) at risk of familial Alzheimer’s disease;^39^ reduced cerebral blood flow can increase SBP;^27^ and cerebral infusion of beta-amyloid (an Alzheimer’s disease pathological hallmark) in rats caused a progressive rise in peripheral BP that was associated with increased levels of the vasoconstrictor endothelin-1 within the brain.^40,41^ Cerebral hypoperfusion is a central and proximal process in the pathophysiology of early Alzheimer’s disease, rather than simply a consequence of diminished metabolic demand. Reversing the reduction (and misdirection) of cerebral blood flow could restore cognitive function, provided synaptic and neuronal damage is not too advanced. Recognising physiological effects of reduced cerebral blood flow, such as elevation of peripheral BP in some people, could play an important part in screening for cerebral vascular dysfunction and identifying those in whom therapeutic interventions might be tested. As more detailed scanning data are collected on UK Biobank participants and as longer-follow up into older age is undertaken, in coming years it will be possible to explore the associations of our instruments for risk of pre-clinical Alzheimer’s disease with cerebral blood flow, and of both of these with subsequent incident Alzheimer’s disease.

DBP was not associated with risk of pre-clinical Alzheimer’s disease when using either the parental dementia instrument score or the participant genetic instrument score, suggesting that DBP may not be elevated due to pre-clinical Alzheimer’s disease. Some studies have suggested a similar DBP-Alzheimer’s disease relationship to SBP, in that elevated mid-life DBP and low late-life DBP is associated with Alzheimer’s disease.^42^ However, DBP has also shown a complex relationship with age where DBP peaks at age 50 due to increased systemic vascular resistance and then declines thereafter.^43^ This is due to large arteries becoming stiffer and therefore having less elastic recoil to support the DBP.

### Study limitations

As noted above we are unable to test the hypothesis fully due to the lack of information on cerebral blood flow. Ideally, we would want a large human study with repeat measures of cerebral blood flow and BP such that the bidirectional hypothesis could be tested further with our instrumental variables. Currently, no such study exists, though the global increase in very large human resources like UK Biobank that increasingly have repeat imaging data, and further follow-up of participants, could make this possible in the future.

The instrumental variables we have used do not allow us to test instrument strength directly and neither do they allow us to quantify the effect of pre-clinical Alzheimer’s disease, because we do not have a measure of the disease at that stage. Thus, we cannot rule out bias due to weak instrument, and importantly we can only indicate the direction of association between the instrumented risk of pre-clinical Alzheimer’s disease and BP. It is not possible to determine whether this is a clinically important magnitude of association. We used available linked primary care and hospital admission data to exclude participants with diagnosed prevalent and 5-year follow-up incident diagnosed Alzheimer’s disease. However, currently and at the time we undertook analyses, although hospital admissions data are available cohort-wide (which include the main reason for hospital admission, plus other underlying health conditions), primary care data were only available for a subset of the cohort (45%). Those who were not linked to GP records are a select group largely from England. Analysis of the subset that have both hospital data and primary care data showed that the age-specific cumulative incidence of dementia is more than halved when primary care data are not included,^44^ as dementia is primarily diagnosed and managed in primary care. This misclassification could have biased our main results by a failure to exclude patients with cognitive impairment or clinical Alzheimer’s disease. We chose a 5-year follow-up *a priori* although recognise that it could be beneficial for future studies to exclude patients who develop dementia over an even longer follow-up period, in addition to obtaining primary care record linkage for the whole cohort, to be able to better isolate pre-clinical disease processes.

In the parental dementia instrument score, we were not able to focus specifically on Alzheimer’s disease as the baseline questionnaire only collected whether the participant’s mother or father had ever suffered from Alzheimer’s disease or dementia. Although Alzheimer’s disease is the most common form of dementia, it accounts for 60-80% of all dementia cases,^45^ which means 20-40% of parental cases that contributed to the score did not have Alzheimer’s disease, which could lead to weak instrument bias. We included biological parents and those who had adopted the UK biobank participant. We did this because the parental dementia instrument should capture environmental factors as well as genetic factors. Of the total 501,420 included in the main analyses, 7,414 (1.48%) had adopted parents, meaning these are unlikely to have had any major impact on the results.

In the participant genetic instrument score, the SNPs had been selected for being associated with Alzheimer’s disease, so this score is perhaps a more specific instrument for risk of pre-clinical Alzheimer’s disease, although it should be noted that most of the diagnoses in GWASs used to establish the participant genetic instrument score were of clinically probable rather than the ‘gold standard’ neuropathologically confirmed Alzheimer’s disease which requires post-mortem examination. However, the positive predictive value of a diagnosis of probable Alzheimer’s disease, made using the criteria used for those GWASs, is >80%,^46^ and the fact that the results are in a consistent direction for both instruments despite differences in relevance/specificity is reassuring.

### Conclusions

Our detailed analyses using two different instrumental variables provides evidence that SBP, but not DBP, is elevated in people at risk of pre-clinical Alzheimer’s disease but without any clinical evidence of cognitive impairment or dementia. Genetic instrumental variable analyses have been previously used to explore risk factors for Alzheimer’s disease, including effects of disrupted sleep, BP and lipid lowering drug targets.^17–25,47,48^ We are also aware of one previous study that compared results from a family based instrument and a genetic instrument in UK biobank data, as we have (in that case to explore intergenerational BMI effects).^49^ Beyond that study and ours, we are not aware that any other studies have compared family based and genetic instrumental variable to address a health question. We have obtained preliminary evidence that pathogenic processes in pre-clinical Alzheimer’s disease increase BP. Obtaining a better understanding of the changing relationship with BP at different stages of Alzheimer’s disease may enable optimisation and targeting of therapies more effectively. Human population studies can use this design in the future to identify pre-clinical disease processes, increase our understanding of the early pathogenic cascades, and inform strategies to delay or prevent Alzheimer’s disease.

## Supporting information

STROBE checklist

## Data Availability

All data produced in the present work are contained in the manuscript

## Acknowledgements

This research has been conducted using the UK Biobank Resource under Application Number 19278.

We thank the International Genomics of Alzheimer’s Project (IGAP) for providing summary results data for these analyses. Full funding information and acknowledgement for IGAP can be found in Kunkle et al.^37^

Dr Alice Carter for providing code for extracting antihypertensive medication use from nurses interview medication use in UK Biobank. Dr Sean Harrison for providing examples of code for Mendelian randomization analysis.

## Contributions

JCP conceptualized the project. JCP and DAL designed the methods with input from EA and EH. JCP completed the analyses, interpreted the findings, and drafted the manuscript, with further input from DL, EH and SL. Review and editing by JCP, EH, EA, SL and DAL.

