## Supplementary Methods, Tables & Figures for "Interrogating the complex relationship between Alzheimer’s disease and blood pressure: two instrumental variable approaches to focus on pre-clinical stages of Alzheimer’s disease in UK Biobank"

**Supplementary material**

**Supplementary Methods**

*Assessment of confounders*

All confounders were measured as part of participants’ in-person assessment and were decided *a priori* from UK Biobank data showcase, before having access to the data. If missing from baseline assessment, we replaced with that recorded at later assessments. If missing from all assessments, we marked as missing. We used a continuous age variable and a binary sex variable. Due to a large amount of missing data for the age participants left education, we used participant’s highest qualification as a measure of education, and categorised into three ordered categories: (1) CSE/O-Level/GCSE or equivalent (~43%); (2) NVQ/HND/HNC/A-levels/AS levels/Other professional qualification or equivalent (~23%); (3) College/University degree or equivalent (~32%). There were 20 categories available for ethnicity, many with sparse data, so we combined into a binary variable of ‘white’ (white, British, Irish, any other white background; ~94%) and ‘aggregated ethnic minority groups’ (African, Asian or Asian British, Bangladeshi, black or black British, Caribbean, Chinese, Indian, mixed, Pakistani, white and Asian, white and black African, white and black Caribbean, any other Asian background, any other black background, any other mixed background, other ethnic group; ~5%). We combined categories of smoking into never (~55%), previous (~35%) and current (~11%) smokers. We kept body mass index (BMI) as a continuous variable. For physical activity we grouped into: no days (~35%); 1-2 days (28%); or 3 or more days (36%) per week of >10 minutes of exercise. We used ‘preference for adding salt to food’ categorised into: never/rarely (~55%); sometimes (~28%); usually (~12%) and always (~5%) as a proxy measure of dietary salt intake. We categorised alcohol into 6 ordered categories of use: never (~8%); special occasions only (~12%); 1-3 times per month (~11%); 1-2 times per week (~26%); 3-4 times per week (23%) and daily/almost daily (~20%). Finally, we used the Townsend deprivation index as a measure of socio-economic position.

**Supplementary Figures**

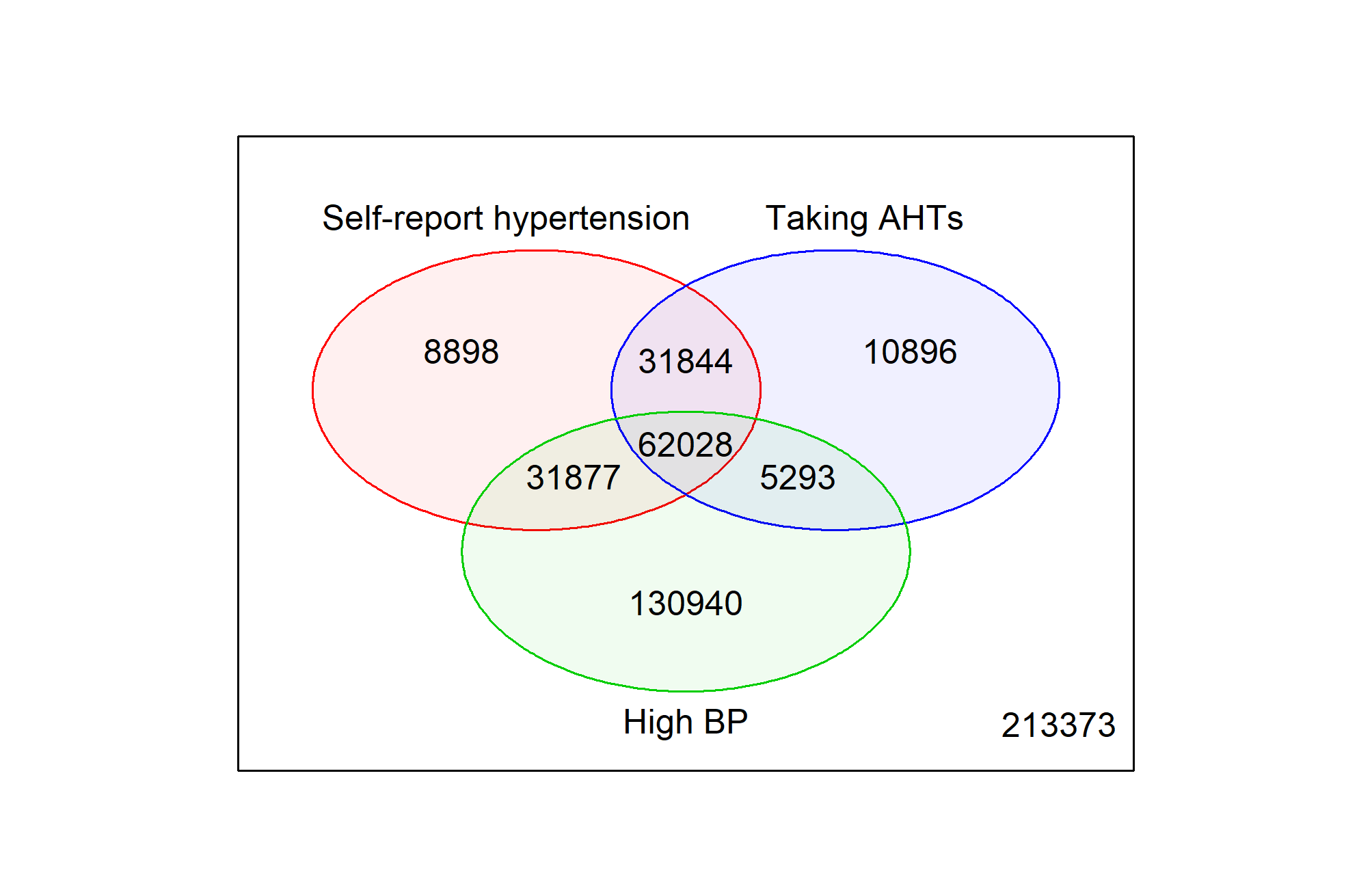

**Supplementary Figure 1** Venn diagram showing composition of Hypertension variable

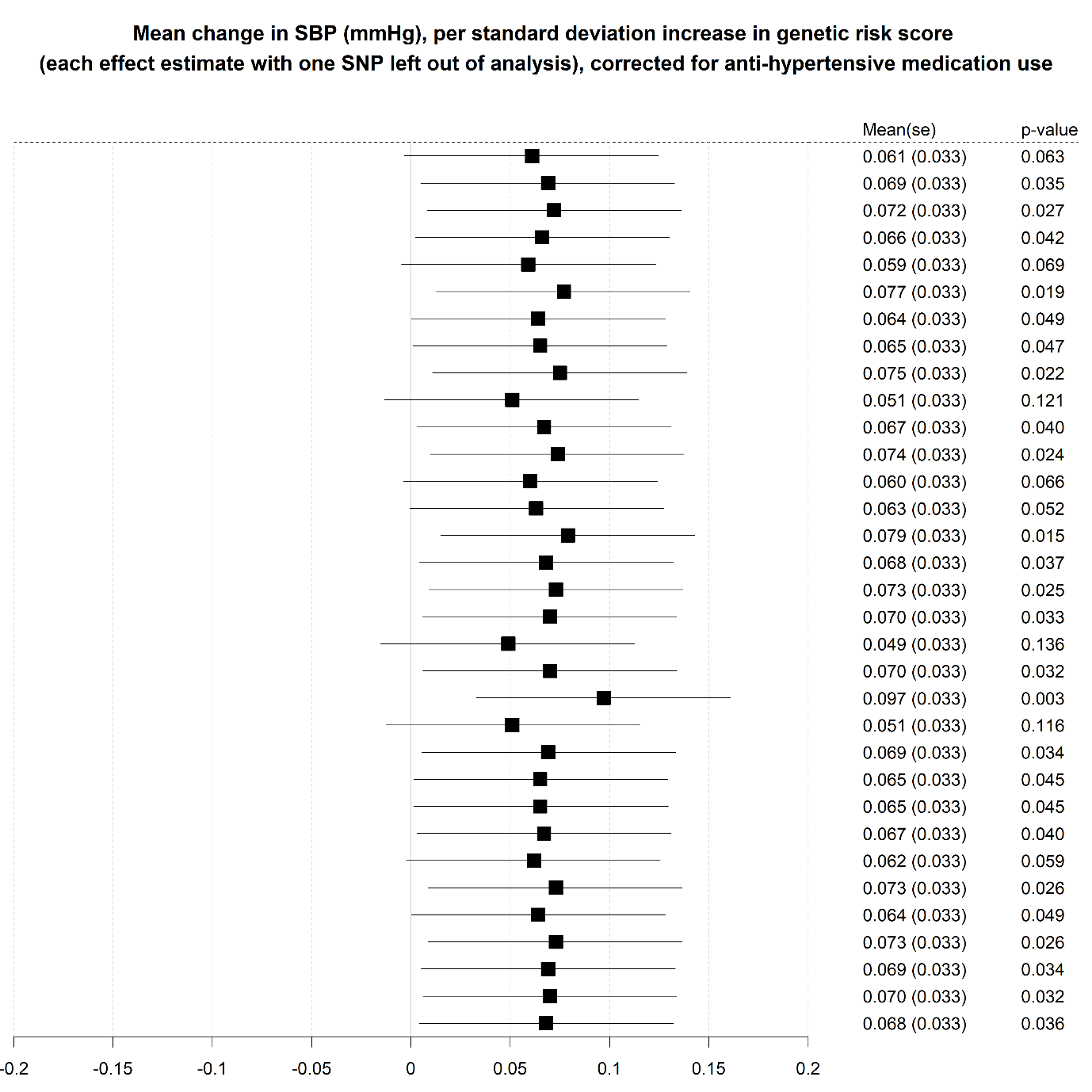

**Supplementary Figure 2** Leave one out analyses of SNPs in the GRS to assess potential for influential outliers in analysis of SBP

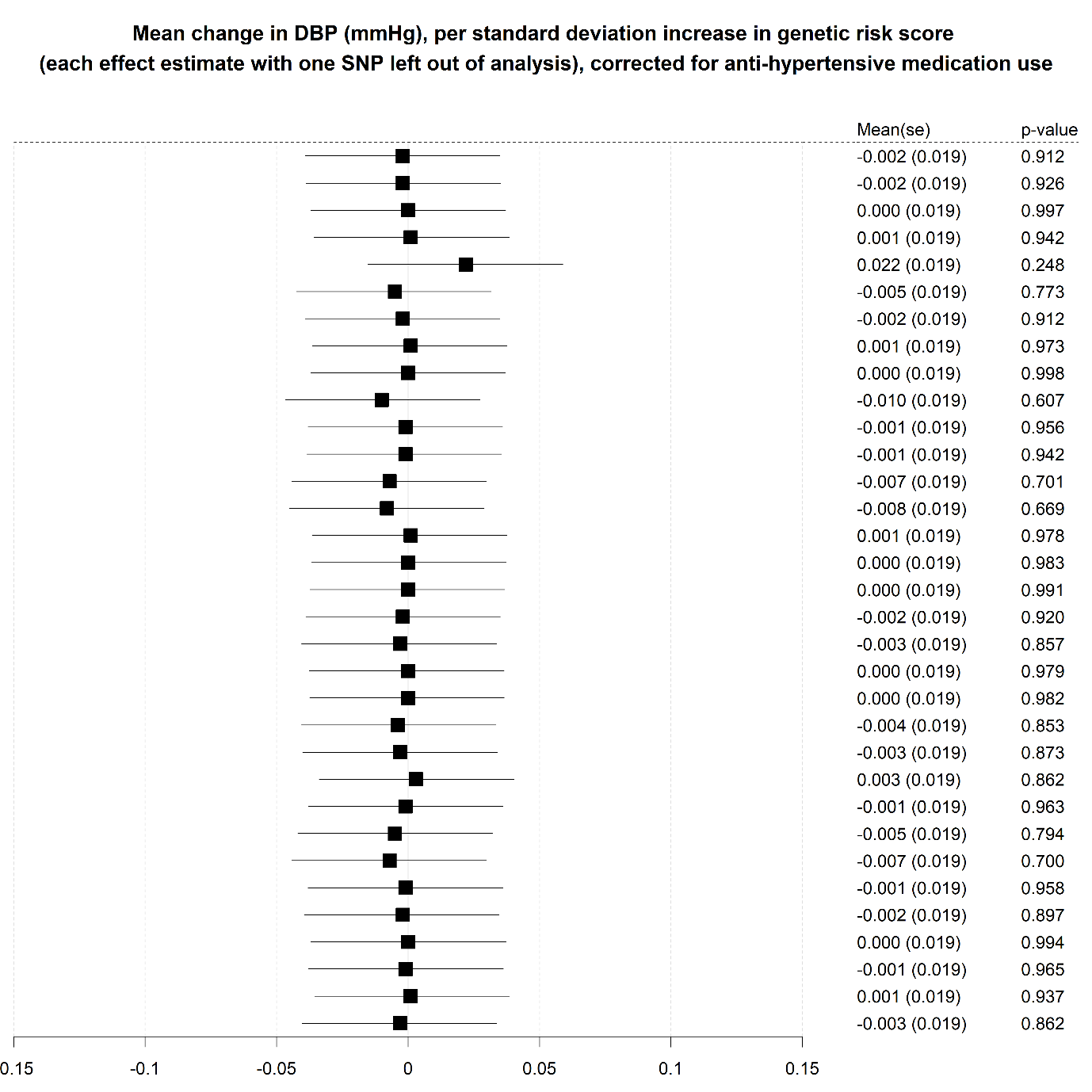

**Supplementary Figure 3** Leave one out analyses of SNPs in the GRS to assess potential for influential outliers in analysis of DBP

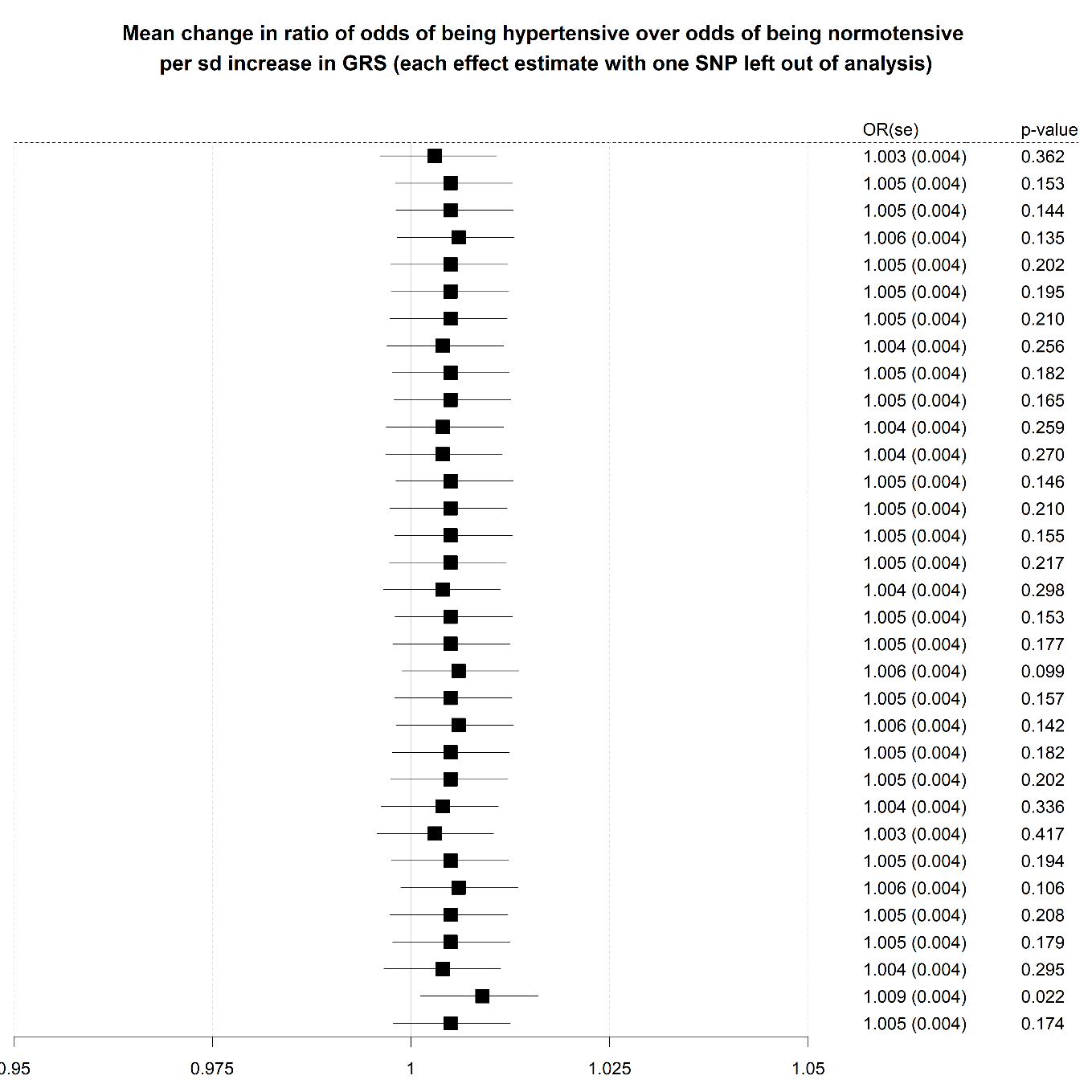

**Supplementary Figure 4** Leave one out analyses of SNPs in the GRS to assess potential for influential outliers in analysis of hypertension

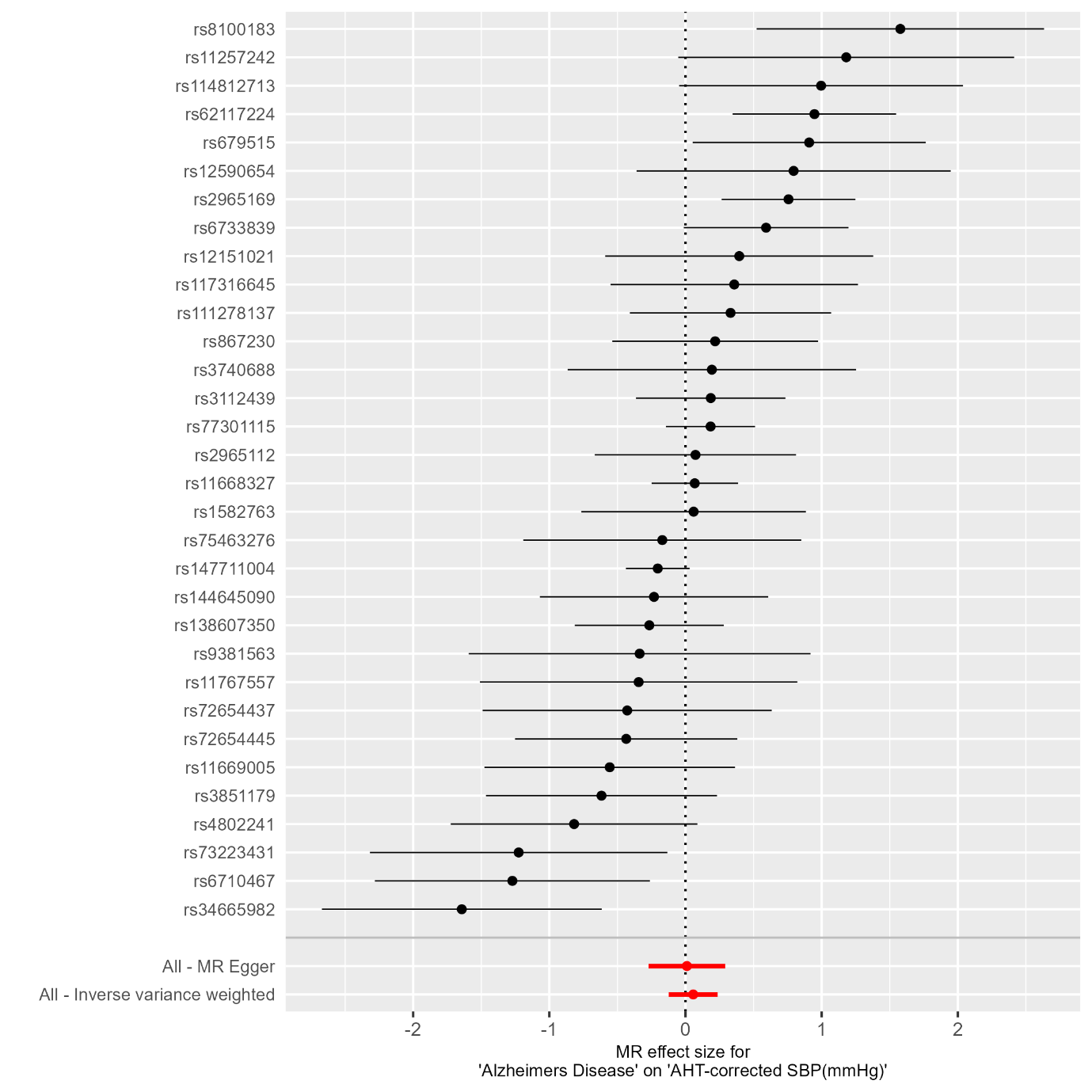

**Supplementary Figure 5** Evaluation of between-SNP heterogeneity. Association of each SNP within the GRS with systolic blood pressure. MR Egger and IVW meta-analyses summaries are shown in red.

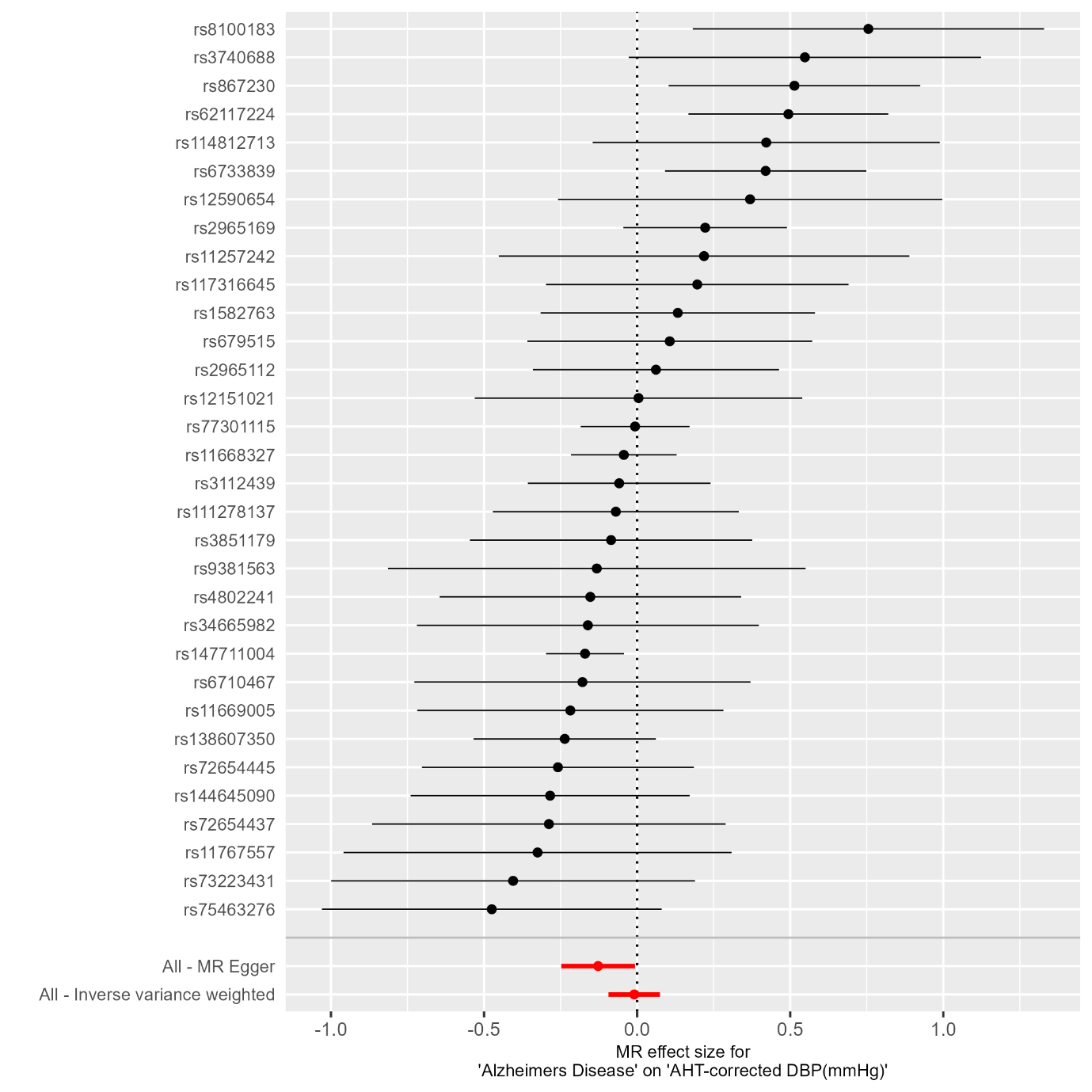

**Supplementary Figure 6** Evaluation of between-SNP heterogeneity. Association of each SNP within the GRS with diastolic blood pressure. MR Egger and IVW meta-analyses summaries are shown in red.

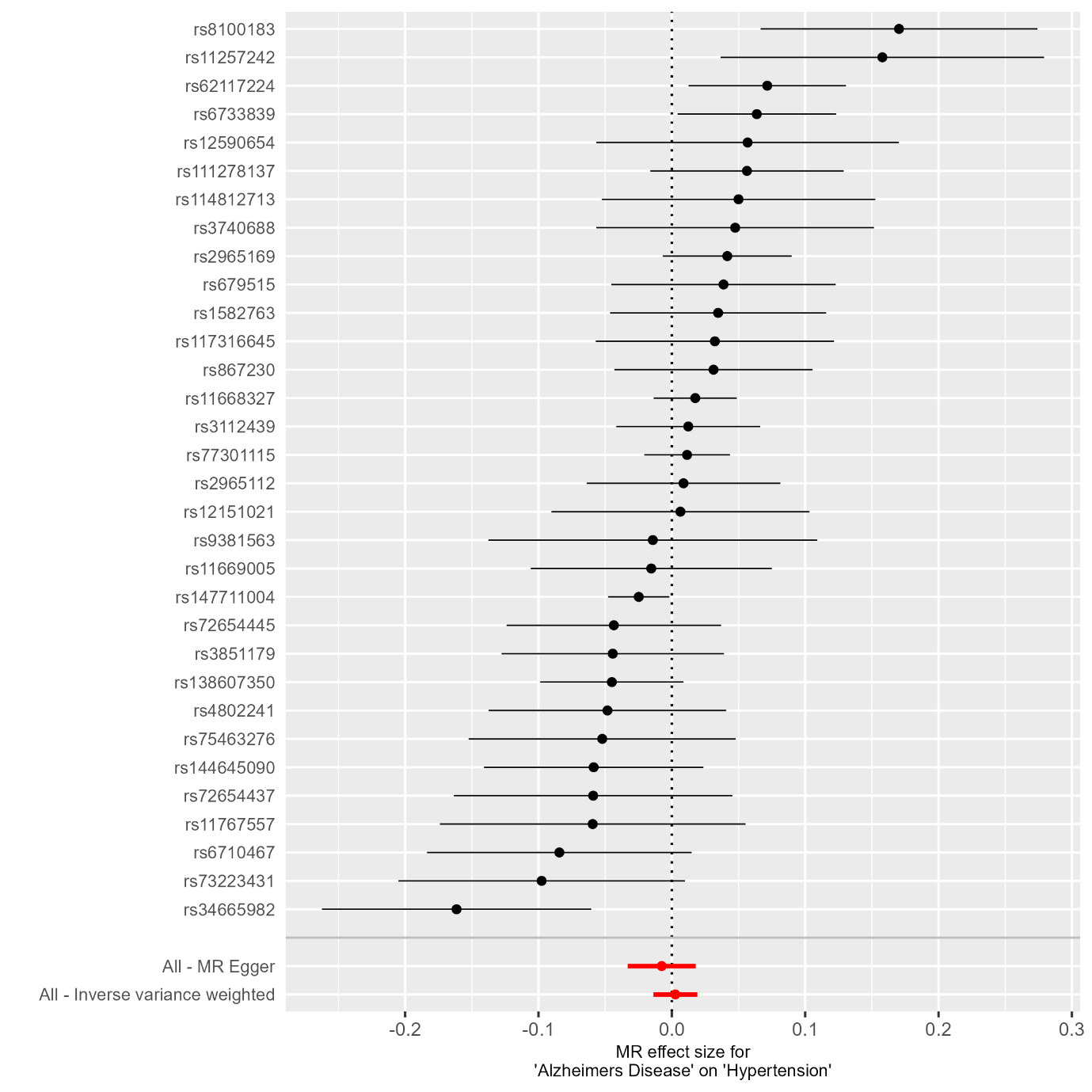

**Supplementary Figure 7** Evaluation of between-SNP heterogeneity. Association of each SNP within the GRS with binary hypertension. MR Egger and IVW meta-analyses summaries are shown in red.

**Supplementary Tables**

**Supplementary Table 1** ICD-9 and ICD-10 codes to extract from Primary Care records and Hospital Episode Statistics

| ICD-9 dementia codes | ICD-10 dementia codes |
| --- | --- |
| 290.2, 290.3, 290.4, 291.2, 294.1, 331.0, 331.1, 331.2, 331.5 | A81.0, F00, F00.0, F00.1, F00.2, F00.9, F01, F01.0, F01.1, F01.2, F01.3, F01.8, F01.9, F02, F02.0, F02.1, F02.2, F02.3, F02.4, F02.8, F03, F05.1, F10.6, G30, G30.0, G30.1, G30.8, G30.9, G31.0, G31.1, G31.8, I67.3 |

**Supplementary Table 2** Missing data

| **Outcome** | **SBP** | | | **DBP** | | | **Hypertension** | | |
| --- | --- | --- | --- | --- | --- | --- | --- | --- | --- |
|  | **n** | **missing** | **% missing** | **n** | **missing** | **% missing** | **n** | **missing** | **% missing** |
| following exclusions, missing parental score and SBP | 445,167 |  |  | 445,168 |  |  | 445,911 |  |  |
| **covariate added** |  |  |  |  |  |  |  |  |  |
| age | 445,167 | 0 | **0.00%** | 445,168 | 0 | **0.00%** | 445,911 | 0 | **0.00%** |
| ses | 444,627 | 540 | **0.12%** | 444,628 | 540 | **0.12%** | 445,368 | 543 | **0.12%** |
| edu(qualifications) | 438,099 | 6,528 | **1.49%** | 438,100 | 6,528 | **1.49%** | 438,800 | 6,568 | **1.50%** |
| ethnicity | 436,845 | 1,254 | **0.29%** | 436,846 | 1,254 | **0.29%** | 437,538 | 1,262 | **0.29%** |
| smoking | 435,505 | 1,340 | **0.31%** | 435,506 | 1,340 | **0.31%** | 436,195 | 1,343 | **0.31%** |
| bmi | 434,019 | 1,486 | **0.34%** | 434,020 | 1,486 | **0.34%** | 434,371 | 1,824 | **0.42%** |
| salt | 433,994 | 25 | **0.01%** | 433,995 | 25 | **0.01%** | 434,346 | 25 | **0.01%** |
| alcohol | 433,764 | 230 | **0.05%** | 433,765 | 230 | **0.05%** | 434,115 | 231 | **0.05%** |
| exercise | 433,764 | 0 | **0.00%** | 433,765 | 0 | **0.00%** | 434,115 | 0 | **0.00%** |
